# Extinction of COVID-19 Clusters in a Lebanese Village: A Quick, Adapted Molecular and Contact tracing

**DOI:** 10.1101/2020.11.28.20240077

**Authors:** Amanda Chamieh, Rania Warrak, Lucie Tawk, Omar Zmerli, Claude Afif, Jean-Marc Rolain, Eid Azar

## Abstract

There is growing evidence of cluster transmission and superspreading of SARS-CoV-2, implying heterogeneous dispersion. We discuss the successful containment of COVID-19 local outbreak in Bcharreh, the small town of 4500 inhabitants, in Northern Lebanon. We look at the dynamics of cluster transmission and viral load evolution throughout the outbreak.

SARS-CoV-2 PCR test was proposed to all exposed individuals. Persons under investigation that tested negative by PCR were periodically retested. We define: a cluster as more than 3 people with a common suspicious or confirmed SARS-CoV-2 positive contact, clinical cure as the resolution of symptoms, and virologic cure as SARS-CoV-2 PCR Cycle threshold(Ct) >35. We analyzed all obtained Ct into corresponding clusters and performed a time series analysis.

A total of 713/871 SARS-CoV-2 PCR tests were performed at Saint George Hospital University Medical Center (SGHUMC) from April 5^th^ 2020 -June 14^th^ 2020. We used the LightMix® Modular SARS-CoV-2 (COVID19) E, N, and RdRP-genes (Tib Molbiol, Berlin, Germany). Week one of epidemiologic surveillance began on March 31^st^ when the first case was detected. A strict lockdown was imposed on Bcharreh village 5 days later, on top of the national lockdown. We identified 4 different clusters ranging from 3 to 27 cases and 3 sporadic unrelated cases.

Almost 70% of each cluster was diagnosed within 7 days. After 2 weeks, we saw a significant increase in the average initial diagnostic Ct 27.9 to 34.72 (P<0.0001). A total of 73/74 SARS-CoV-2 PCR positive individuals achieved cure (98.6%). We recorded one death of a 90-year-old man with multiple comorbidities.

In explosive new epidemics, we can derive from previous experience and not be blinded by it. To safely navigate out of the lockdown, focus on where new transmission is likely to emerge and accordingly target available diagnostic technologies.

## Introduction

The COVID-19 pandemic overwhelmed healthcare systems worldwide with thousands of cases diagnosed daily(1). We all became acquainted with the basic reproduction number Ro, which is a calculation of the average number of secondary transmissions caused by a single primary case in a fully susceptible population(2).

According to Lloyd-Smith(3) and the London School of Hygiene and Tropical Medicine COVID-19 Working Group(4), a population estimate of Ro would underestimate the high degree of individual level variation or overdispersion in the distribution of COVID-19. Based on the globally estimated Ro of 2-3(5,6), the overdispersion parameter *k* was estimated at 0.1. This suggests that 80% of secondary transmission is caused by around 10% of infectious individuals. In Hong Kong, 20% of cases were responsible for 80% of the local transmission, and *k* was estimated at around 0.45 (7). Some countries such as Iran, Italy, Spain and France saw a devastating surge of locally transmitted cases where epidemiologic link was no longer traceable(8). Other countries that also saw imported cases of COVID-19 had much less transmissions than expected if the Ro of 2-3 was accurate.

This growing evidence in support of cluster transmission and superspreading of SARS-CoV-2 implies that dispersion of the disease is heterogeneous. An analysis of Japanese contact tracing of COVID-19 confirmed patients identified 61 clusters, with clusters reaching more than 100 individuals, related to only 22 index cases (9). Thus, early outbreaks models should account for the proportion of cases that do not transmit as well as Ro to properly estimate individual variation in infectivity.

According to known outbreak report models, the number of expected cases in Lebanon, from February 21^st^ when the first imported case was confirmed, was 600,000 individuals within 2-3 months and at least 1500 deaths(10). This did not reflect the observed reality of cluster transmission. Lebanon is a small country of 10,452 km^2^ with 6.9 million inhabitants with tightly knit societies that played a major role in successful contact tracing. We discuss the successful containment of COVID-19 local outbreak in Bcharreh, the small town of 4500 inhabitants, in Northern Lebanon. We look at the dynamics of cluster transmission and viral load evolution throughout the outbreak.

## The Study

In Lebanon, the first diagnosed case in February 21 2020 led to a gradual lockdown of the country, prohibiting all gatherings by mid-March 2020. According to Ministry of Public Health (MOPH), since SARS-CoV-2 was declared a Public Health Emergency of International Concern (PHEIC), Lebanon built a capacity for 576 hospital beds, 234 ICU beds and 263 ventilators.

One of the pillars of Lebanese Response Plan was mobilizing municipalities, mayors and creating local resources and networks for contact tracing. Disparity of healthcare system is apparent with many underserviced public governmental hospitals and major university hospital centers located in center of Beirut(11). Luckily, in the construct of Lebanese society, this implies that a community or municipality is totally traceable. The urgency of the situation created an initiative for small villages to perform thorough contact tracing and show up for testing. The citizens of this town waived privacy concerns and the contact tracing was transparent.

SARS-CoV-2 PCR testing was divided among different public and private testing centers in an effort to rapidly test and contain all persons under investigation. The major proportion of testing was conducted at our Saint George Hospital University Medical Center (SGHUMC) Clinical Laboratories starting April 5^th^ 2020 until June 14^th^ 2020.

We use the LightMix® Modular SARS-CoV-2 (COVID19) E, N, and RdRP-genes (Tib Molbiol, Berlin, Germany) as previously described(12). The cycle number threshold values (Ct) is a reflection of quantifiable SARS-CoV-2. A higher Ct confers a lower quantity of SARS-CoV-2.

Week one of epidemiologic surveillance began on March 31^st^ when the first case was detected. By April 5^th^, another 10 cases appeared. A decision was taken by the local corona response team to perform a very thorough contact tracing. A strict lockdown was imposed on Bcharreh 5 days later, on top of the national lockdown. This meant that neither entry nor exit of the town was allowed, ensuring no new imported cases, and that all commerce was shut down.

We proposed performing a PCR-based test on all the exposed individuals and their families regardless of symptoms development. All the individuals were screened and followed up daily. Each positive individual had at least one follow-up PCR test. We continued periodic screening and testing for individuals with positive contacts yet tested negative.

We define: (1) a cluster as more than 3 people with exposure to a common suspicious or confirmed SARS-CoV-2 positive contact, (2) clinical cure as the resolution of symptoms, (3) virologic cure as obtaining a SARS-CoV-2 PCR with a Ct>35, implying no infectivity (13,14). We analyzed retrospectively all obtained Ct into their corresponding clusters and performed a time series analysis.

## Results

A total of 871 PCR tests were performed on the traced contacts of the town of Bcharreh, with 713 in our laboratories at SGHUMC. A total of 74 individuals were SARS-CoV-2 PCR positive. Seventy-three out of seventy-four achieved clinical and virologic cure, at 98.6%. Results at SGHUMC are available for 67 out of 74 diagnosed. There was only one death of a 90-year-old man with multiple comorbidities.

We managed to diagnose 65% (43/67) of the COVID-19 cases in the first epidemiologic week. In week 2, 4 new cases diagnosed (5.9%). In week 3, 9 new cases (13.4%). In week 4, 1 new case (1.49%). In week 5, 7 cases diagnosed (10.4%). In week 6, 1 case diagnosed (1.49%). In week 7, 2 new cases diagnosed (2.9%).

The average initial diagnostic cycle threshold (iCt) gradually increases throughout the study period. We start at an iCt of 27.5 in weeks 1 and 2. In week 3, the average iCt increases to 30.7. In week 4-5, the average iCt increases to 34.25. In weeks 6-7 the average iCt is at 32. The time series analysis showed a significantly increasing trend of iCt, and specifically beyond the date of April 22, 2020, in epidemiologic week 3, from which the average Ct shifts from 27.9 to 34.72 (p<0.0001) (Figure 1).

**Figure 1.**
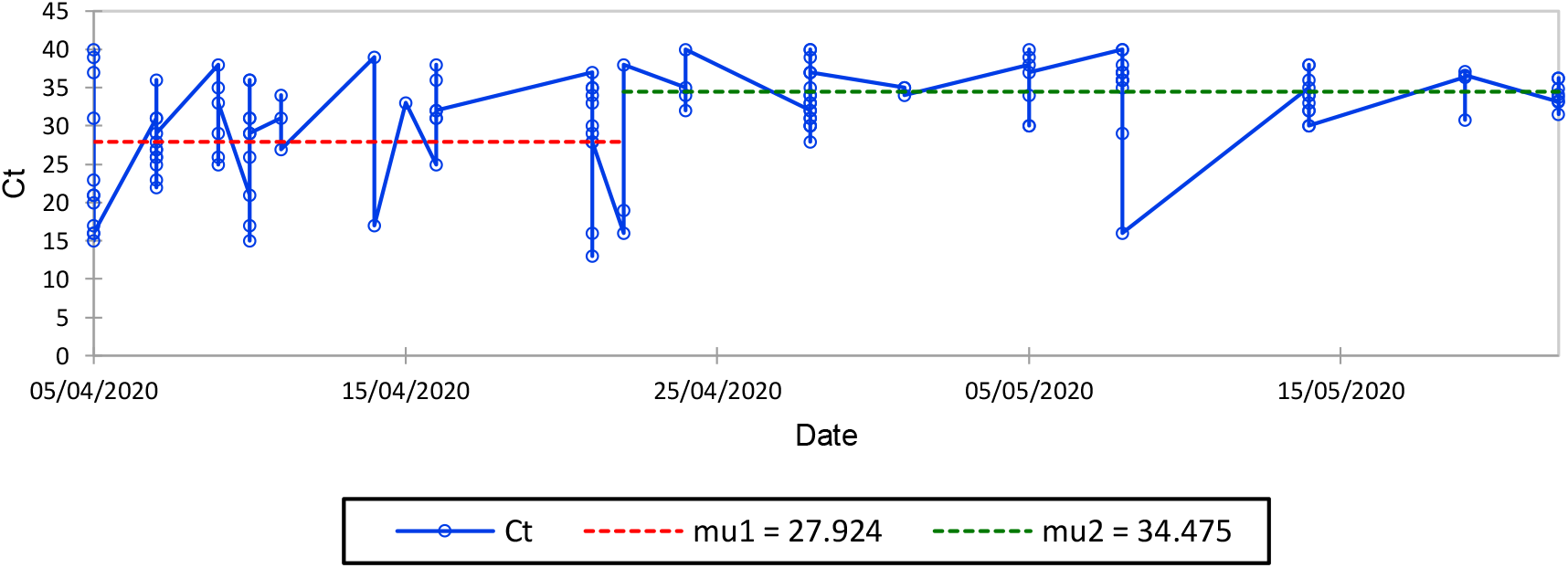
Initial Diagnostic Average Ct per week.

**Figure 2:**
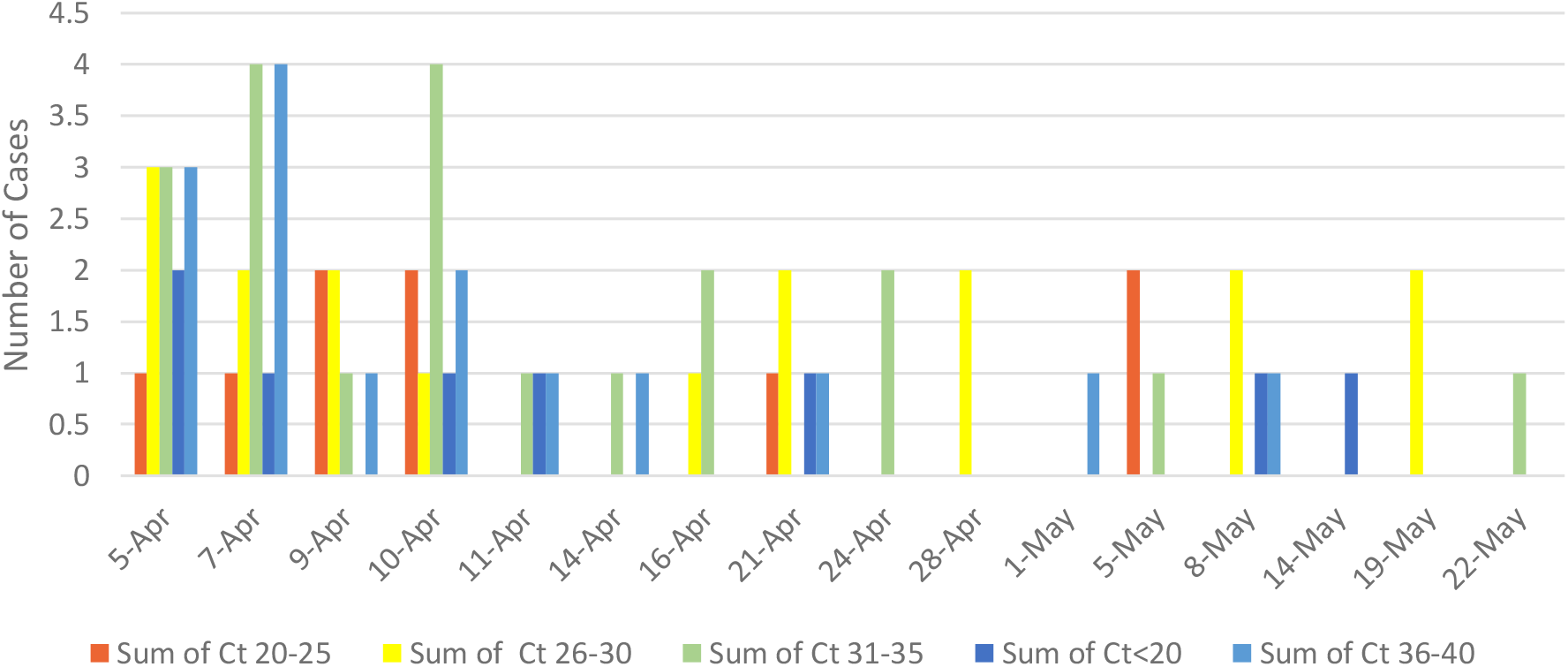
Evolution of Initial Diagnostic SARS-CoV-2 PCR Cycle Number(Ct) Values Throughout Bcharre Outbreak.

## Cluster Transmission

Following the contact tracing that was performed we identified 4 different clusters and a group of sporadic unrelated cases. Three sporadic cases appeared in weeks 2, 4 and 7, at iCt of 17, 35, and 31, respectively. The cluster sizes ranged from 3 to 27 cases. In week 1, 3 clusters were traced, H, T and L. A cluster G appeared in week 3 with possible epidemiologic linkage to clusters L and H.

Cluster H is related to a common healthcare related exposure, totaling 8 cases of which 5/8 (63%) were diagnosed in week 1. In week 2, 2 cases were diagnosed while in week 3 the last one. Cluster H had an average iCt of 29.8 in week 1, which increased to 34 in week 2 and 3. No statistically significant trend was detected in this cluster.

Cluster T is one main family exposure with different households that had one index case. This cluster totaled 27 cases, of which 19/27(70%) were diagnosed within 1 week. In week 2, there was one new case at an average Ct of 31. We diagnosed 3 cases in each of weeks 3 and 5 at an average iCt of 31.7 and 35, respectively. In week 7, one case was diagnosed at iCt of 33. Time series analysis showed a significantly increasing trend of average iCt (p<0.00001).

Cluster L is 4 different intertwined families with different households with the same common exposed individuals. This cluster totaled 24 cases, of which 19/24 (79%) were diagnosed in week 1 at an average iCt of 29. In week 3, 1 case was diagnosed at an iCt of 35. In week 5, 3 cases were diagnosed at an average iCt of 32. In week 6, the last case was diagnosed at an average iCt of 32. Time series analysis did not detect any trend in this cluster.

Cluster G is a family with relations to cluster L and contact with cluster H yet epidemiologic linkage was no longer fully traceable. This cluster is composed of 5 individuals, 4 (80%) of which were diagnosed in epidemiologic week 3 at an average iCt of 22.25. In the first week of appearance of this cluster, 80% of the cases were diagnosed. The last case was diagnosed in week 5 at iCt of 16. There was no trend detected in this series.

## Discussion/Interpretations

When a novel pathogen is encountered, we do not yet know the role of individual variation on transmission. This may explain why we observe a significant discrepancy between reality and the mathematical model based on global population-wide estimation of Ro. Following the example of the Chinese government, several countries opted for extreme social distancing, national and international lockdown, and quarantine of suspicious and confirmed cases. Soon enough, the world was in a global lockdown as an attempt to stop the spread of COVID-19(15,16).

Containment of cases in small countries with smaller towns and villages appeared to be easier in some communes such as Ferrera Erbognone in the Province of Pavia in the Italian region Lombardy, located about 45 km southwest of Milan, and Zahara de La Sierra in southern Spain. There are around 1500-2000 inhabitants in both these towns. They applied a strict lockdown and recorded zero cases of COVID-19(17,18).

This is similar to our described case of Bcharreh where 4 clusters were closely followed, and all contacts were traced molecularly. The investigative, intrusive nature of neighborly interactions facilitated a smooth and comprehensive contact tracing and subsequent SARS-CoV-2 PCR testing of all contacts, resulting in extinction of all COVID-19 cluster within 8 weeks.

The common limitation in facing emerging pathogens such as SARS-CoV-2 is the unknown epidemiologic characteristics and transmissibility that may impede and complicate effective control strategies. More than often, this leads to delays in proper management and an overwhelmed healthcare system. The various proposed mathematical models and projections may be misleading and far from real life empiric experience. Most models for SARS-CoV-2 were based on assumptions from the 1918 Flu pandemic, which we know follows a homogenous transmission with a dispersion factor *k* of 1. This means that clusters had very little role in the 1918 Flu pandemic, contradicting the transmission pattern of the current COVID-19 pandemic(3,19).

We also know that the higher the proportion of non-transmitting individuals, i.e. the more over dispersed, the more likely that disease extinction occurs and the chain of transmission dies out. However, if an outbreak does result, it is much more explosive than predicted by Ro. So, a disease with high over dispersion that does not result in an outbreak probably lacked the superspreading event. This is where lockdown and social distancing come in. In addition, this may explain why the introduction of SARS-CoV-2 in France dating back to December 2019 did not result in an outbreak at that time(20).

After 2 weeks, we saw a significant decrease in the average initial diagnostic Ct (P<0.0001) probably affected by the lockdown, and preventive measures such as mask use, social distancing and hand hygiene. The decreasing initial diagnostic viral load means that the identified case probably has a lower burden of disease and is expected to be less contagious. The highest transmission occurs in the first few days before and after symptom onset(21), but we also know that around 50% of infections are asymptomatic(22,23). In addition, the duration of infectivity was thought to last as long as viral shedding continues early on in the pandemic. We now know that after 11 days, the risk of transmission is negligeable (13,14,24–26).All this contributes to individual viral load variation and infectivity. A decreasing viral load either indicates an infection with a very low inoculum after lockdown and cluster containment or a recovering illness.

The rapid diagnosis and strict follow up of each emerging cluster within its first week likely contributed to the containment. We managed to diagnose an average 70% of each cluster within the first 7 days. By the end of second week, the cluster was almost extinct. Despite the low Ct indicating a high viral load in cluster G, this cluster died off and did not lead to further cases. This was more likely a sub cluster from cluster L, hence the nonuniformity and variation in Ct of cluster L. We believe the drop in average iCt may signify the appearance of a new case with ability to super spread hence creating an alarm to test and isolate.

Cluster T had 4 cases with a very low average iCt of 16 in 4 individuals on day 1 of surveillance, yet did not yield any further subclusters. On the other hand, cluster L had 2 cases with a very low average Ct of 16 in 2 individuals on day 6 of surveillance. This appearance of a low Ct further along the timeline from outbreak onset may have contributed to the subclusters in cluster L. On the other hand, cluster G, with an average Ct of 17.5 in 2 individuals on 17 of surveillance and 1 individual with Ct 16 on day 34 of surveillance, did not result in any other cases. This highlights the heterogeneity of transmission in clusters and how viral load impact is variable depending on the measures taken.

Moreover, the cut-off value for a negative PCR differs between laboratories, manufacturers, and genes tested. A cut-off was suggested at Ct>35 because above that yields no viable culture hence no transmissibility(13,27), whereas another found that at Ct>24 there is no viable culturable viral RNA(14). Clinically, this is important for treatment of individual cases. However, epidemiologically, any still traceable amount of DNA is an added, necessary tool for contact tracing. Upon a novel pathogen outbreak, aggressive contact tracing allows for a 3-fold more effective targeted control as compared to random control in outbreak containment(3). It is more thorough than relying on symptom development since a high proportion will not demonstrate any and may be subjective(22,28). This is unrelated to clinical status of a patient and is more informative on a larger scale from a public health and epidemiologic point of view. Thus, we suggest a strict molecular contact tracing as a valid method of detecting more cases.

In our work, we focused mainly on the obtained SARS-CoV-2 PCR results and on contact tracing details. It was not possible at the time to account for individual genetic polymorphisms nor genetic predispositions suggested by blood group types. Moreover, to date, the most accurate tool remains whole genome sequencing to detect clonal outbreaks. This may have added to the precision of following the evolution of COVID-19 outbreak in our country.

In explosive new epidemics, we must derive from previous experience and not be blinded by it. In novel emerging pathogens, anticipating heterogeneity in transmission should be used to plan disease control programs and risk-stratify populations for public health interventions. Prompt and targeted public health interventions prevent and mitigate superspreading events and pave the way for a timely interruption of the chain of transmission during containment. In turn, this reduces the disruption of healthcare services and society(19). This allows localized, targeted lockdowns and avoids stunting the whole society on a national and international scale. Performing aggressive molecular contact tracing, using average initial diagnostic Ct values, and analyzing subsequently the whole genome sequence of SARS-CoV-2 over time can help us implement the necessary interventions.

## Data Availability

Data available upon request

